# Basic helix-loop-helix transcription factor *BHLHE22* monoallelic and biallelic variants cause a neurodevelopmental disorder with agenesis of the corpus callosum, intellectual disability, tone and movement abnormalities

**DOI:** 10.1101/2024.10.11.24312856

**Authors:** Carolyn Le, Emanuela Argilli, Elizabeth George, Tuğba Kalaycı, Zehra Oya Uyguner, Birsen Karaman, Tanju Demirören, Stephanie DiTroia, Delphine Heron, Isabelle Sabatier, Lance H. Rodan, Katta Mohan Girisha, Periyasamy Radhakrishnan, Carol Saunders, Bonnie Sullivan, Emily Fleming, Javeria Raza Alvi, Tipu Sultan, Henry Houlden, Stephanie Efthymiou, Maria J. Guillen Sacoto, Melanie Goodman, Lucie Pierron, Jean-Madeleine De Sainte-Agathe, Alexandra Durr, Elliott H. Sherr

## Abstract

*BHLHE22* encodes a Class II basic helix-loop-helix transcription factor (bHLH). It is expressed exclusively in the retina and central nervous system (CNS), and functions as an important regulator of retinogenesis and neuronal differentiation. Mice lacking bhlhe22 show nearly complete loss of three brain comminsure, including the corpus callosum. Here we report eleven individuals from nine unrelated families with *BHLHE22* variants, with a neurodevelopmental disorder presenting with absent or limited speech, severely impaired motor abilities, intellectual disability (ID), involuntary movements, autistic traits with stereotypies, abnormal muscle tone. The majority of individuals have partial or complete agenesis of the corpus callosum (ACC). Additional symptoms comprised of epilepsy, variable dysmorphic features, and eye anomalies. One additional individual had spastic paraplegia without delayed development and ACC, expanding the phenotype to milder and later onset forms. Four individuals carry *de novo* missense variants within the highly conserved helix-loop-helix domain while seven individuals from five unrelated families carry a recurrent homozygous frameshift variant, p.(Gly74Alafs*18). Our findings implicate *BHLHE22* variants in causing a previously unidentified autosomal dominant and recessive neurodevelopmental disorder associated with ACC, severe motor, language, and cognitive delays, abnormal tone, and involuntary movements. To our knowledge, this is the first report of Class II bHLH variants in humans shown to significantly disrupt brain development, cognition, and movement.

## INTRODUCTION

The corpus callosum is the largest interhemispheric commissural tract with nearly 200 million axons traversing the cerebral midline and functions to transmit information between the hemispheres, including sensory, motor, and cognitive signals (Aboitiz and Montiel, 2003).

Corpus callosum dysgenesis (CCD) is a condition that results from the disruption of the early stages of fetal cortical and callosal development, including corpus callosum hypoplasia (hCC) and partial or complete ACC. It has been recently reported to be as common as 1 in every 2,000 births (Hofman et al, 2020) and has been linked to varying degrees of neurodevelopmental, cognitive, and behavioral issues (Paul, 2011). In 30-55% of individuals with CCD, the etiology has been identified to be genetic (Edwards et al, 2014, Sajan et al, 2013) and while over 800 single genes have been associated with CCD, many hundreds remain to be discovered.

With this goal, through our institution, we recruited and performed trio exome sequencing (proband and both biological parents) on a large cohort of research participants (n=504), who have been diagnosed with CCD. Within the cohort, we identified an index case with a *de novo* variant in the gene *BHLHE22*, not reported in the gnomAD database and predicted to be damaging, p.(Glu251Gln). Through international collaborations, we subsequently recruited ten additional individuals who had similar clinical features and either heterozygous and *de novo* or biallelic inherited variants in *BHLHE22*.

The gene *BHLHE22* (previously called *BHLHB5*) maps to cytogenetic locus 8q12.3. It encodes a putative 381-amino acid protein with a molecular weight of 36.9 kD from a single exon, which includes an N-terminal proline-rich domain, a glycine-rich domain, a polyglycine-serine region, a helix-loop-helix (HLH) domain, and a C-terminal alanine-rich region (Xu et al, 2002). BHLHE22 belongs to the basic helix-loop-helix family of transcription factors, characterized by two alpha-helices that mediate dimerization, and an active domain that directs deoxyribonucleic acid (DNA) binding to the E-box (Bertrand et al, 2002). This superfamily of dimeric transcriptional regulators is an important promoter of cell fate determination, proliferation, and differentiation in the development of the nervous system (Massari & Murre, 2000). The bHLH proteins have been broadly classified based on tissue distribution. Class I bHLH proteins are ubiquitously expressed and bind exclusively to the E-box site. Class II bHLH proteins show tissue-specific expression (Bertrand et al., 2002), and include members of the NeuroD, Nscl, and Olig families (Dennis et al, 2019). Within the Olig family, BHLHE22 is expressed exclusively in the CNS and retina (Xu et al, 2002). It is required for the differentiation of neurons in several CNS domains, including the dorsal horn of the spinal cord (Ross et al, 2010), the dorsal cochlear nucleus in the brainstem (Cai et al, 2016), and retinal amacrine cells (Feng et al, 2006). Notably, the loss of *bhlhe22* in the dorsal telencephalon of mice led to nearly complete loss of three fiber tracts crucial for bilateral connectivity of the cerebral hemispheres: the corpus callosum, hippocampal commissure, and anterior commissure (Ross et al, 2012).

In our *BHLHE22* cohort, four individuals carry heterozygous *de novo* missense variants in the HLH domain, and seven individuals carry recurrent homozygous frameshift variants. 3/4 individuals with heterozygous variants present with partial or complete ACC, motor deficits, hypotonia, limited speech, ID, autistic traits with stereotypies, and oculomotor/vision issues. In addition to these features, the majority of individuals with the recurrent homozygous variant are unable to acquire speech, have microcephaly, lower limb or appendicular spasticity, and are non-ambulatory, frequently with dystonia and/or dyskinesia. Additional features seen in some individuals include seizures, scoliosis, non-specific facial dysmorphic findings, and feeding difficulties. Interestingly, one heterozygous carriers of a de novo missense variant exhibited spastic paraplegia solely and therefore adding *BHLHE22* to the long list of genes involved in spastic paraparesis (Darios et al, 2021). The clinical and magnetic resonance imaging (MRI) findings in the *BHLHE22* cohort align with a previously unidentified neurodevelopmental disorder with structural brain malformations.

## SUBJECTS AND METHODS

### Subjects

Individual 1 was initially enrolled in the Disorders of Cerebral Development: A Phenotypic and Genetic Analysis Study at the Brain Development Research Program of the University of California, San Francisco (UCSF) as part of a larger cohort study of participants with confirmed corpus callosum abnormalities. This study protocol was approved by the UCSF Institutional Review Board (IRB) and the enrolled subject had informed consent provided by the parent/guardian.

Individuals 2-11 were identified through MatchMaker Exchange and informed consent was obtained by their respective institutions (Sobreira et al, 2015). Available medical records and brain imaging scans/reports were collected from each individual and reviewed by the UCSF research team, including a pediatric neuroradiologist and pediatric neurologist.

### Sequencing

Sample identity quality assurance checks were performed on all samples. Exome or genome sequencing was performed using Illumina platforms (NextSeq 500, Illumina NovaSeq 6000, Illumina NovaSeqX, or Illumina HiSeq 4000). Individuals 6 and 7 were tested through the genetic testing platform GeneDx (Guillen Sacoto et al, 2020). Bioinformatics analyses were performed according to the best practices of GATK (V.3.4). Variant filtering was based on minor allele frequency <0.1% for autosomal dominant (AD) searches and <1% for autosomal recessive (AR) searches, and a predicted deleterious impact on the gene/protein for all samples.

Bidirectional sequences were assembled, aligned to reference gene sequences based on human genome build GRCh37/hg19 or GRCh38/hg38. For individuals 1, 3, 9, 10, and 11, sequence validation and segregation analysis for the candidate variants were performed through Sanger sequencing.

## RESULTS

Our work, supported by a series of domestic and international collaborations, identified eleven individuals with heterozygous *de novo* or biallelic homozygous variants in the gene *BHLHE22*. Clinical, imaging, and genetic findings of the complete cohort are described in Table 1. A schematic representation of the variant locations, tolerance landscape plot, and protein sequence of human *BHLHE22* and its vertebrae orthologs as well as brain MRIs are outlined in Figure 1 and Figure 2, respectively.

**Figure 1.**
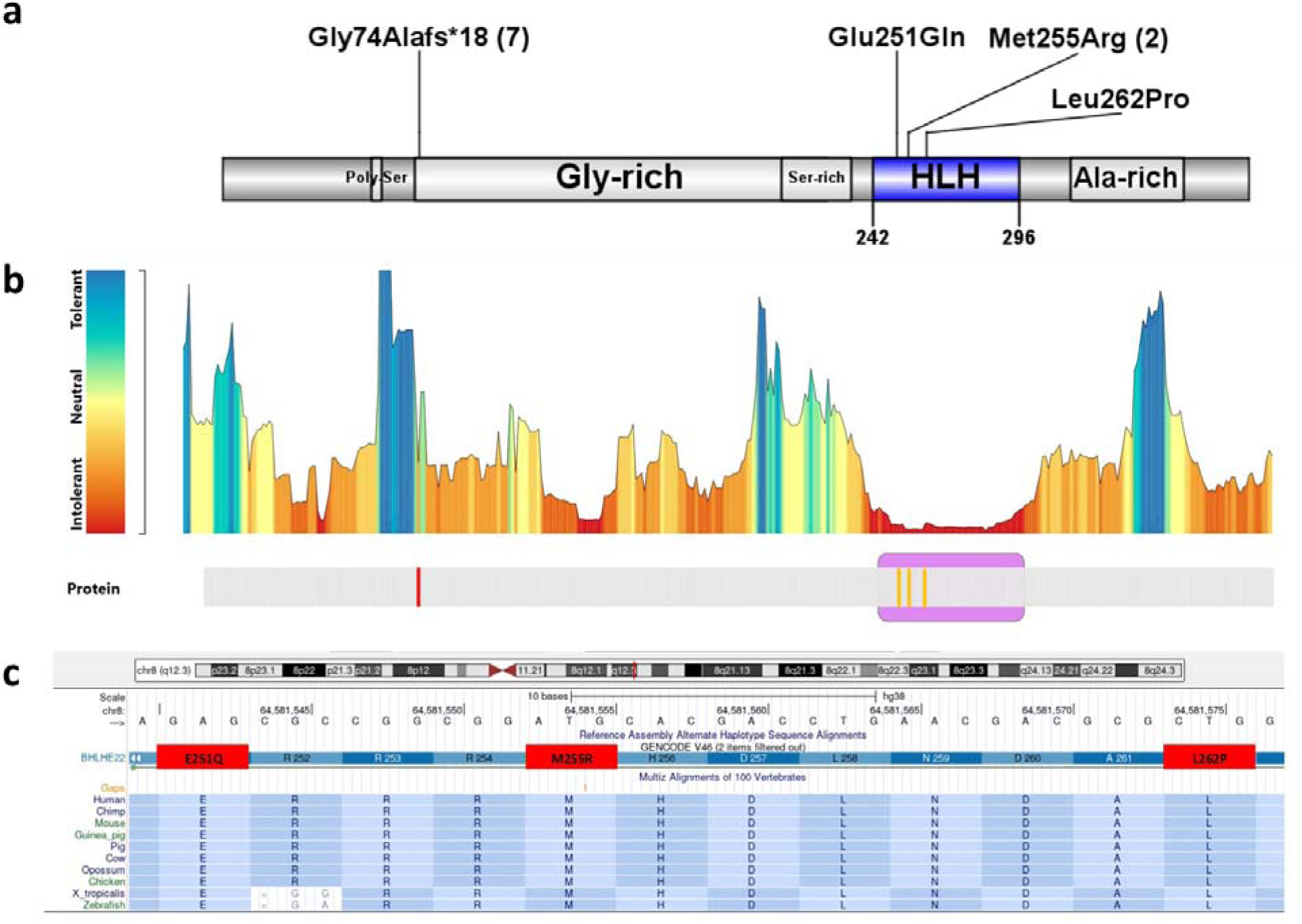
A. Schematic representation of the BHLHE22 protein with the described *de novo* p.(Glu251Gln), recurrent p.(Met255Arg), and p.(Leu262Pro) variant both contained in the HLH active domain important for dimerization, and biallelic recurrent variant p.(Gly74Alafs*18). B. Tolerance landscape plot of the BHLHE22 protein provided by the MetaDome web server (https://stuart.radboudumc.nl/metadome/). The tool identifies regions of low tolerance to missense variations based on local non-synonymous over synonymous variants ratio from gnomAD. The *de novo* variants in our cohort are contained in intolerant/highly intolerant regions (in red) of the landscape. C. Various sequence alignment provided by the University of California, Santa Cruz genome browser (https://genome.ucsc.edu/) show the protein sequence of the human BHLHE22 protein and its orthologs in different vertebrate species with the mutated residues in red.

**Figure 2.**
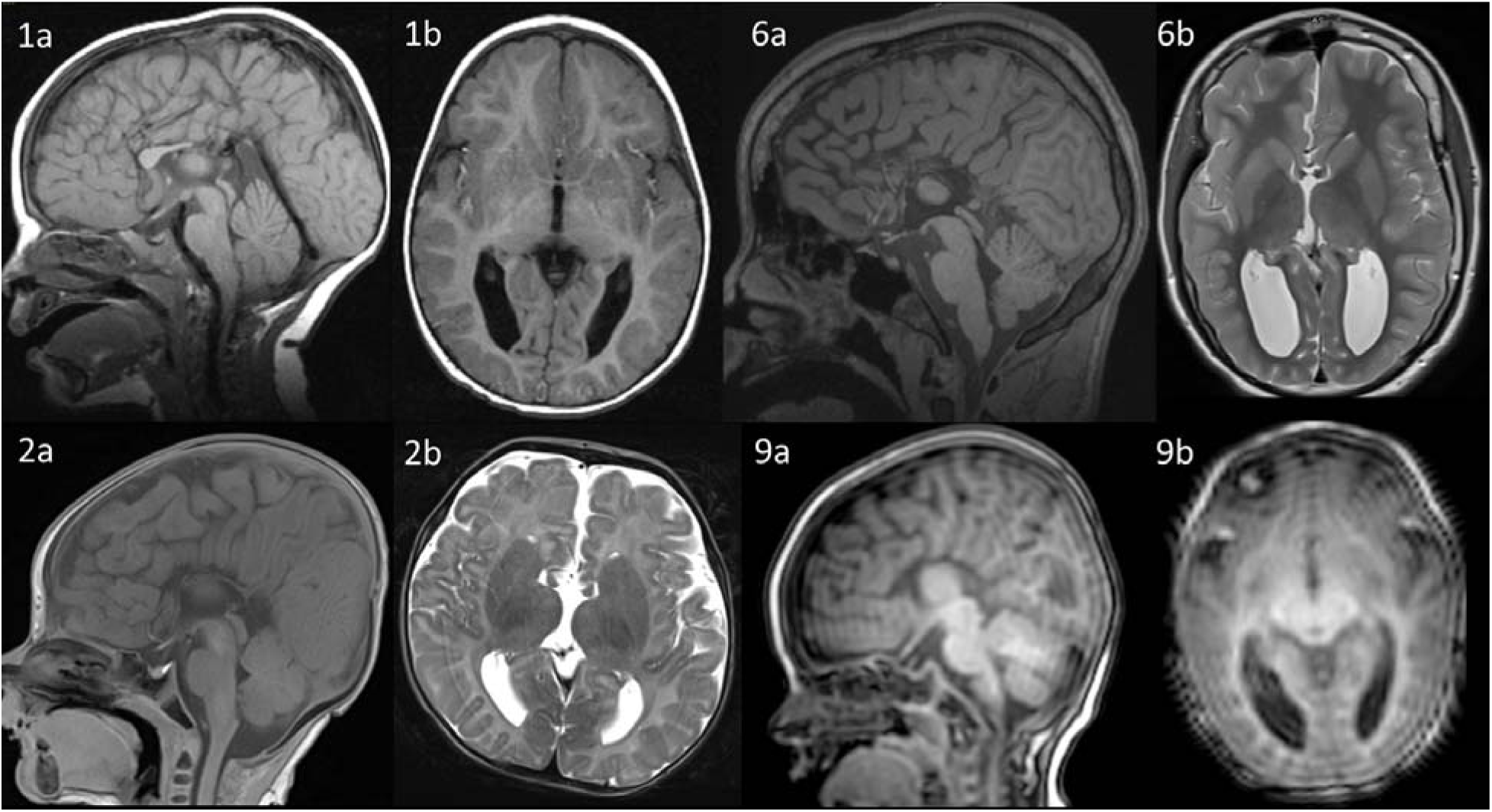
Brain imaging of patients in the *BHLHE22* cohort. Individual 1’s T1 sagittal (1a) indicate partial ACC with absence of the rostrum, posterior body, and splenium. T1 axial scan (1b) shows colpocephaly. Individual 2’s T1 sagittal (2a) shows complete ACC and T2 axial (2b) shows colpocephaly. Individual 6’s T1 sagittal (6a) and T2 axial (6b) shows complete ACC and colpocephaly. Individual 9’s T1 sagittal (9a) shows complete ACC and T1 axial (9b) shows colpocephaly.

**Table 1.**
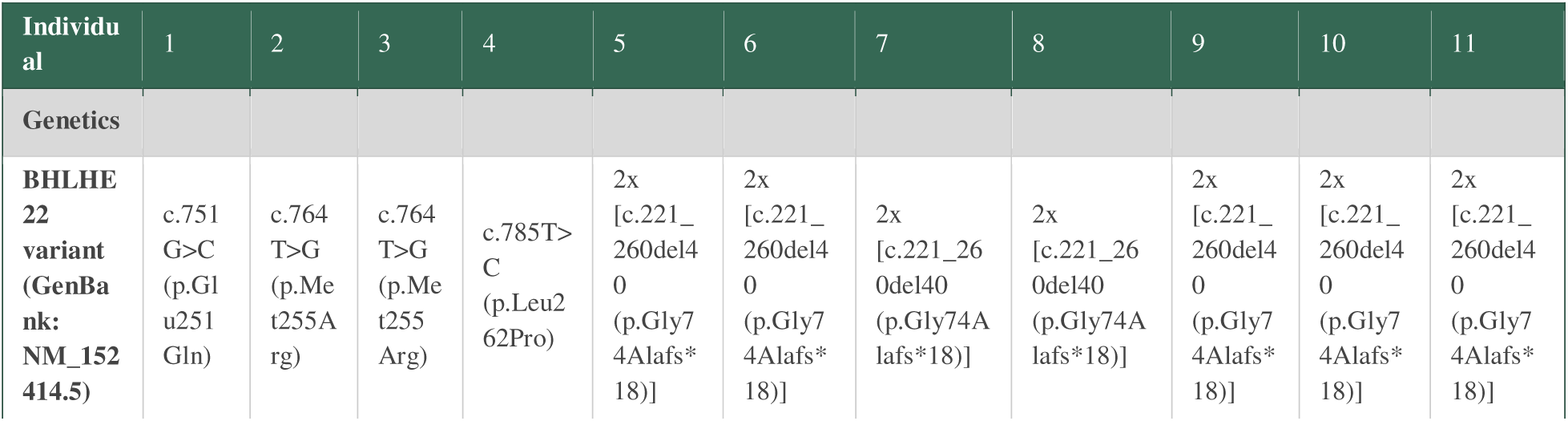
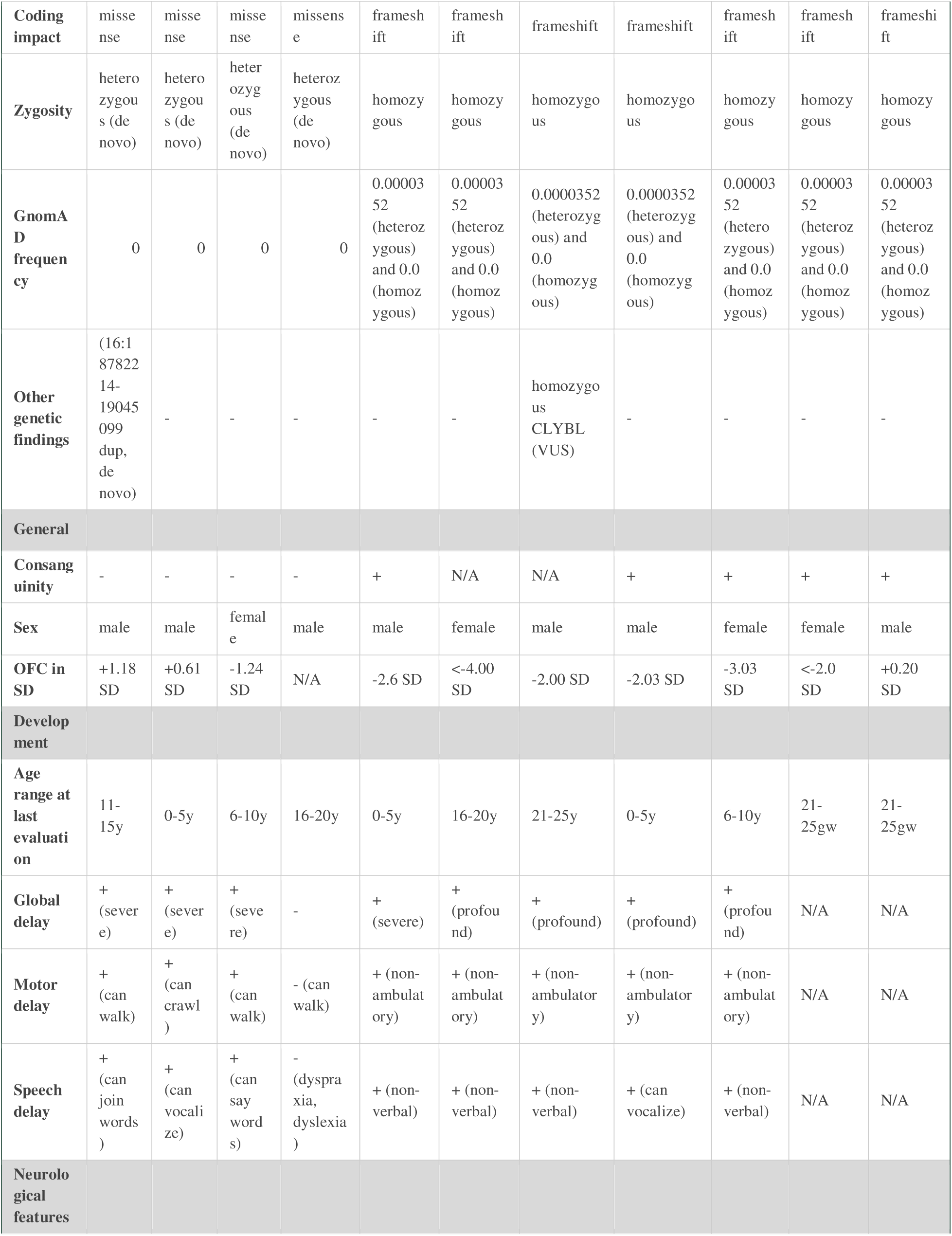
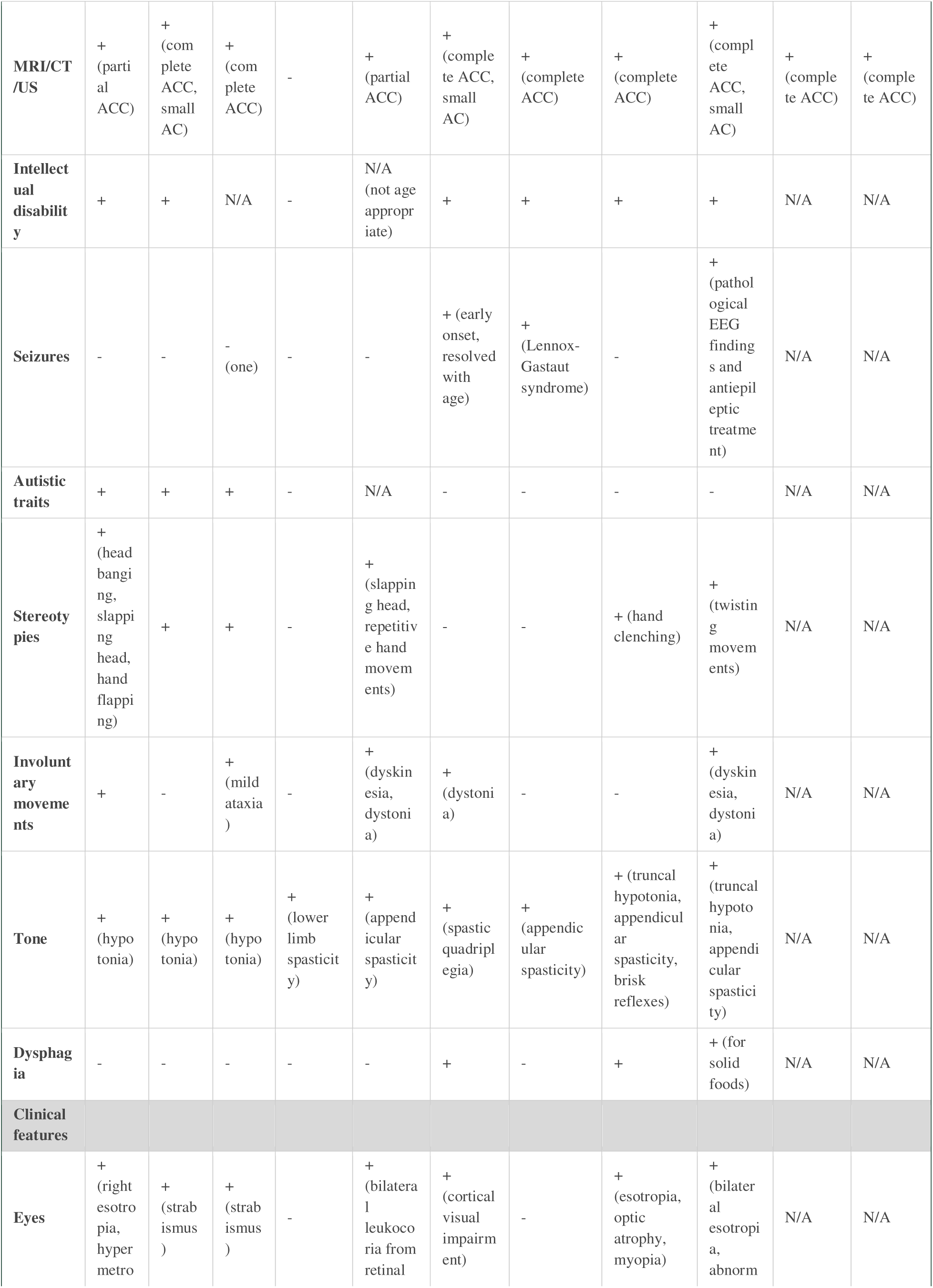
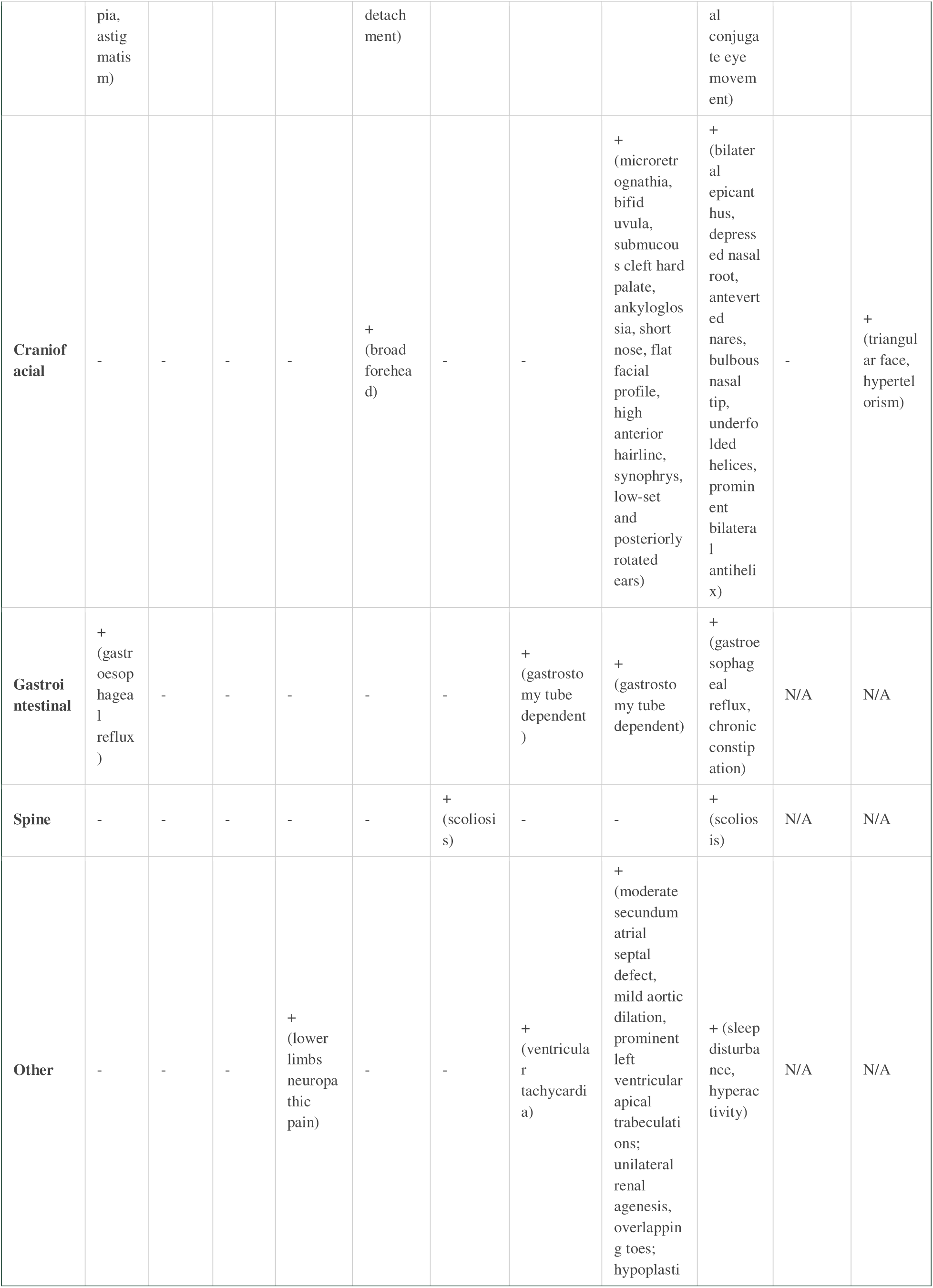
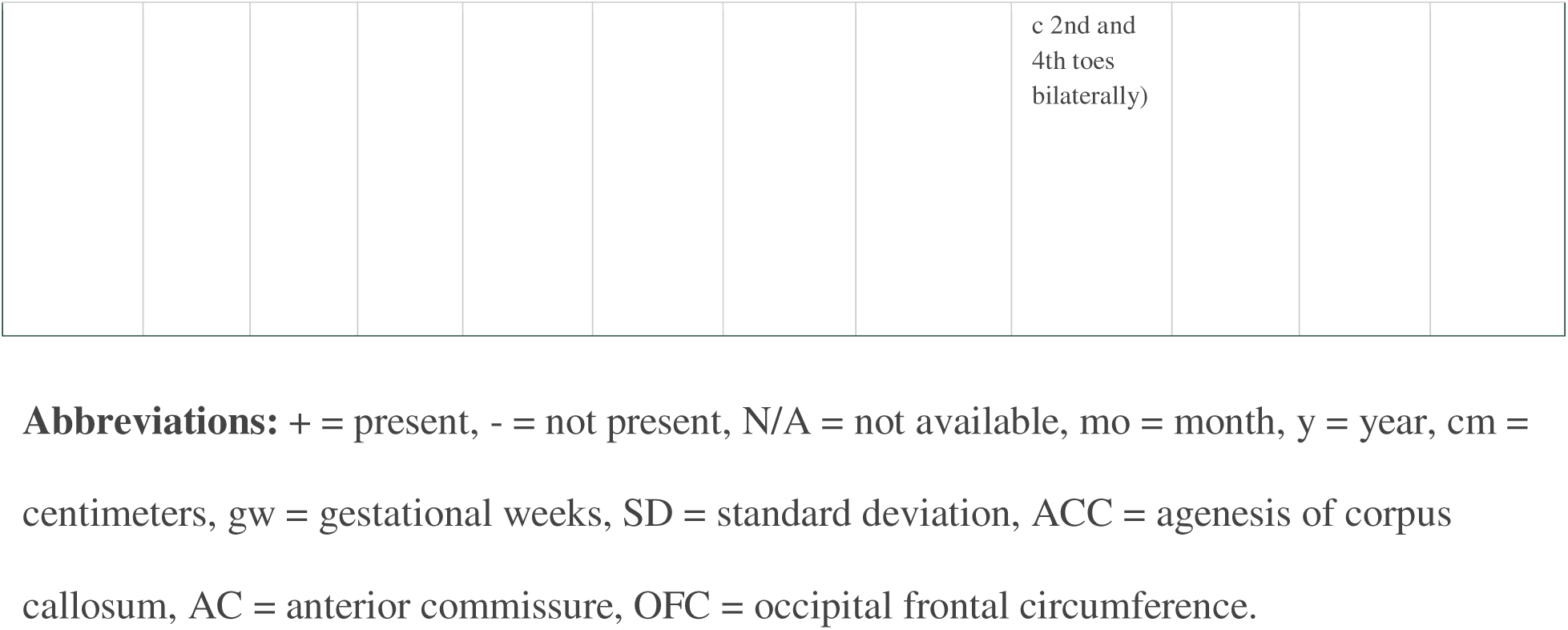

### Individual 1

Individual 1 is a 11-15 years old male born at term via elective cesarean section to non-consanguineous parents. His birth weight and length were within the normal range. His neonatal period included feeding complications and acid reflux. A brain MRI was performed for early developmental concerns. It revealed partial ACC with absence of the rostrum, posterior body, and splenium, as well as colpocephalic configuration of the occipital horns of the lateral ventricles. The anterior commissure is present while the hippocampal commissure is absent and the overall white matter volume is mildly diminished (Figure 2). An electroencephalogram (EEG) was performed and was not revealing. His childhood development was characterized by severe motor, speech, and social delay. Walking was moderately delayed, but achieved and he is able to join words. Between 6-10 years old, he met the Diagnostic and Statistical Manual of Mental Disorders, 4th Edition, Text Revision (DSM-IV-TR) criteria for autism spectrum disorder. Other neurological findings include central hypotonia, repetitive self-stimulatory, and self-injurious movements such as slapping his head, hand flapping, and head banging.

Oculomotor and vision issues include right esotropia, astigmatism, and significant hyperopia. Family history was negative for epilepsy, cerebral palsy, and ID. Trio exome sequencing (ES) revealed a *de novo* missense variant in the gene *BHLHE22,* NM_152414.5: c.751G>C, p.(Glu251Gln). This variant is absent from gnomAD, has a CADD-phred score of 31, and is located in the highly conserved helix-loop-helix domain. Another genetic finding was a *de novo* 263 kb microduplication Chr16:18782214-19045099, affecting seven genes including ARL6IP, associated with spastic paraplegia 61, an autosomal recessive disorder. A second hit to suggest pathogenicity of this variant in ARL6IP was not found.

### Individual 2

Individual 2 is a 0-5 years-old male whose prenatal brain imaging showed complete ACC. He was born at term with normal birth weight, length, and OFC. His neonatal history included slow weight gain and hypotonia. A brain MRI in infancy confirmed complete ACC with a small anterior commissure and no other brain malformations (Figure 2). The hippocampal commissure was not observed. At his most recent examination, he presented with severe motor, speech, and social delay. He is unable to walk, but can crawl. He can vocalize, but has no intentional words. He shows autistic traits including stereotypies and problems with social interaction. His additional medical history includes poor eye tracking and strabismus.

Chromosome microarray and karyotype analysis were normal. Trio ES detected a *de novo* missense variant in the gene *BHLHE22,* NM_152414.5: c.764T>G, p.(Met255Arg). This variant is absent in gnomAD, has a CADD-phred score of 31, and is located in the highly conserved helix-loop-helix domain.

### Individual 3

Individual 3 is a 6-10 years-old female born to non-consanguineous parents at term. In the neonatal period, she had a brain MRI that revealed complete ACC. In infancy, she had an isolated episode of generalized tonic clonic seizure. She had developmental concerns including speech delay and severe gross motor delays, but walking was achieved. Neurological examination revealed ataxia, mild hypotonia, and autistic traits. Other clinical history includes strabismus. Chromosomal microarray was negative. Clinical trio ES was performed revealing a *de novo* variant in *BHLHE22,* NM_152414.5: c.764T>G, p.(Met255Arg), identical to the variant in Individual 2.

### Individual 4

Individual 4 is an 16-20 years-old male born after an uneventful pregnancy to non-consanguineous parents. The delivery and neonatal period were normal. His psychomotor development was normal. He had some difficulties with concentration, dyslexia, and dyspraxia, for which he had speech therapy and psychomotricity during childhood. He could follow a normal education. In early adolescence, he presented with pain in the lower limb quickly followed by claw toes and lower limbs spasticity. A brain MRI was performed and was not revealing. The corpus callosum was normal. In late adolescence, he had moderate lower limb spastic paraparesis without muscle weakness. The extensive neurological examination was otherwise normal. Family history was unremarkable for any neurological disease. Trio genome sequencing (GS) revealed a *de novo* missense variant in the gene *BHLHE22*, NM_152414.5: c.785T>C, p.(Leu262Pro). This variant is absent from gnomAD, has a CADD-phred score of 32, and is located in the highly conserved helix-loop-helix domain. No other pathogenic or likely pathogenic variants were identified.

### Individual 5

Individual 5 was a 0-5 years-old male born from consanguineous parents. He was born via elective cesarean section. As a toddler, he exhibited truncal hypertonia and limb spasticity with abnormal movements including dystonia, choreiform movements, and stereotypical hand movements. A computed tomography (CT) scan revealed partial ACC with anterior remnant present and colpocephaly. The anterior and hippocampal commissure was not visualized on a limited scan. His development was characterized by severe gross and fine motor delay as he was unable to sit or hold his neck without support. He could hold objects with his hands, but not transfer them. He had no speech development. He experienced bilateral leukocoria from suspected retinal detachment. The retinal detachment was corrected via successful surgery and as his vision improved, he began to smile and play. His most recent evaluation revealed a broad forehead and mild microcephaly (-2.6 SD). Unfortunately, he passed away at the age of 0-5 years.

ES revealed a homozygous frameshift deletion in the gene *BHLHE22,* NM_152414.5: c.221_260del40, p.(Gly74Alafs*18). This variant has been reported in gnomAD only in the heterozygous form with an allele frequency of 3.52e-5 (Chen et al, 2022). Individual 5’s parents and two unaffected siblings are heterozygous carriers, confirmed by Sanger sequencing.

### Individual 6

Individual 6 is a 16-20 years-old female born prematurely at 32 weeks. She was hospitalized in the neonatal intensive care unit due to prematurity. In infancy, seizures began, but resolved by early childhood. An EEG revealed occipital spikes. A brain MRI revealed complete ACC with associated colpocephaly and a small anterior commissure (Figure 2). The hippocampal commissure was not visualized. She exhibits severe motor and speech delay along with severe ID. She is non-verbal and non-ambuluatory with spastic quadriplegia and dystonia in her upper extremities, requiring medications and surgery. She has very limited social interaction and is unable to point due to her dystonia and cortical visual impairment. Other medical issues include thoracolumbar scoliosis and feeding issues with dysphagia. Her most recent growth parameters revealed normal weight and height, while OFC indicated severe microcephaly (<- 4.00 SD). Biological family history was not available. A chromosome microarray revealed multiple regions of homozygosity. ES revealed a homozygous frameshift deletion in the gene *BHLHE22,* NM_152414.5: c.221_260del40, p.(Gly74Alafs*18), identical to Individual 5.

### Individual 7

Individual 7 is a 21-25 years-old male. In infancy, he started to have generalized tonic clonic seizures and was subsequently diagnosed with Lennox-Gastaut Syndrome. He has severe motor delay and is wheel-chair bound with spasticity of all four extremities. He is non-verbal and diagnosed with ID. A brain MRI report indicated complete ACC with colpocephaly.

He has ventricular tachycardia and supraventricular ectopy. He is gastrostomy tube dependent. His most recent growth parameters include normal weight and mild microcephaly (approximately-2.00 SD for his age). His family history is significant for a brother diagnosed with complete ACC and ID. Unfortunately, the brother passed away at 6-10 years-old and genetic testing was not completed. Individual 7 had a normal chromosome microarray, while trio ES revealed homozygous frameshift variants in the gene *BHLHE22,* NM_152414.5: c.221_260del40, p.(Gly74Alafs*18), identical to Individuals 5 and 6. The sequencing results also show a homozygous variant of unknown significance (VUS) in the gene *CLYBL* and a paternally inherited heterozygous pathogenic variant in the gene *ACSF3*, associated with an autosomal recessive disorder that includes neurodevelopmental delays and seizures. A second hit on the gene that would suggest pathogenicity was not found.

### Individual 8

Individual 8 is a 0-5 years-old male to consanguineous parents with 6.5% regions of homozygosity. During infancy, he had feeding difficulties and generalized failure to thrive. A brain MRI report indicated complete ACC with no other malformations. He exhibits severe motor delay as he cannot roll. He can grasp objects with both hands, but does not transfer them. He exhibits severe speech delay as he can vocalize, but has no intentional words. His tone is characterized by truncal hypotonia and peripheral spasticity with brisk reflexes and hand clenching movements.

Oculomotor and vision issues include occasional esotropia, optic nerve atrophy, and high myopia. His eyes do not track, but he can focus on an object briefly. Comprehensive medical history includes gastrostomy tube dependence and oropharyngeal dysphagia. An echocardiogram revealed a moderate secundum atrial septal defect, prominent left ventricular apical trabeculations, and mild dilatation of the ascending aorta. He has unilateral renal agenesis.

Craniofacial feature include microretrognathia, bifid uvula, submucous cleft hard palate, ankyloglossia, short nose, flat facial profile with low facial tone, high anterior hairline, synophrys, and low-set, posteriorly rotated ears. In addition, he has overlapping toes and hypoplastic toes bilaterally. His most recent growth parameters revealed mild microcephaly (OFC of-2.03 SD). ES revealed homozygous frameshift variants in *BHLHE22,* NM_152414.5: c.216_255del40, p.(Gly74Alafs*18), identical to Individuals 5, 6, and 7. ES was performed on the proband and the mother, who is heterozygous for the variant. The father was not available to participate.

### Individual 9

Individual 9 is a 6-10 years-old female and firstborn of first-degree cousins. During pregnancy, ultrasound (US) imaging revealed complete ACC with no other structural anomalies detected. She was born preterm via cesarean section due to rupture of membranes. Birth measurements included normal weight and length. An EEG revealed independent focal epileptiform activities in the right and left frontocentrotemporal regions. She has no clinically recognizable seizures. However, anticonvulsant therapy was initiated. A brain MRI confirmed complete ACC, a small anterior commissure, absent hippocampal commissure, slightly enlarged ventricles with colpocephaly, and moderately diminished white matter volume (Figure 2). She was hospitalized for febrile convulsions secondary to pneumonia. A subsequent EEG displayed a consistent pattern compared to the previous one and her anticonvulsant therapy was changed to a different medication. She showed profound motor delay and did not have head control until early childhood. At the most recent evaluation, she remains unable to sit without support, walk, or point at objects. She is unable to say single words and maintains limited eye contact. She is diagnosed with ID. She has truncal hypotonia, mild spasticity in the lower extremities, dystonia in the arms with dyskinetic movements, mild spasms, and lack of purposeful hand movements.

Oculomotor issues are present with bilateral esotropia and disconjugate eye movement. Her medical history includes gastreoesophageal reflux disease in infancy, chronic constipation, and dysphagia with solid foods. Additionally, she has scoliosis, sleep disturbance, and hyperactivity. She has significant facial dysmorphism including bilateral epicanthus, a depressed nasal root, anteverted nares, a bulbous nasal tip, under folded helices, and prominent antihelices. Her most recent growth parameters show moderate microcephaly (OFC of-3.03 SD).

ES followed by confirmatory Sanger sequencing validation revealed homozygous frameshift variants in *BHLHE22,* NM_152414.5: c.221_260del40, p.(Gly74Alafs*18), identical to Individuals 5, 6, 7, and 8. Both the mother and the father carried this variant in one allele. The asymptomatic mother of Individual 9 also carried a common heterozygous non-frameshift insertion variant of c.667_672dup, p.(Ser223_Gly224dup) in *BHLHE22*, which is not found in any other family members. This additional variant has been reported in gnomAD with an allele frequency of 0.0962.

Family history for Individual 9 is notable for two siblings (Individuals 10 and 11 described below) with ACC, which was discovered in utero. In addition, there was a recent pregnancy, where a prenatal US revealed a normal corpus callosum. This pregnancy progressed without complications, leading to term birth via cesarean section. At a recent examination, the infant displayed normal development. Sanger sequencing revealed a heterozygous frameshift variant in *BHLHE22*, NM_152414.5, c.221_260del40, p.(Gly74Alafs*18). The family’s pedigree is outlined in Supplementary Figure 1 and their Sanger sequencing is displayed in Supplementary Figure 2.

### Individuals 10 & 11

Individual 9’s family history includes two terminated fetal siblings. Individuals 10 and 11 underwent termination both at 20-25 weeks, following the identification of complete ACC and colpocephaly through prenatal US. Other findings include an OFC SD of about <-2.00 and +0.20, respectively. Individual 10 had hypertelorism and a triangular face. Individual 10 and Individual 11 had the same recurrent homozygous variants in *BHLHE22,* NM_152414.5: c.221_260del40, p.(Gly74Alafs*18), identical to Individuals 5, 6, 7, 8, and 9.

## DISCUSSION

We identified eleven individuals from nine unrelated families with monoallelic or biallelic *BHLHE22* variants associated with mild to severe neurodevelopmental delay, most often ACC, and abnormal tone and movements. All four heterozygous *de novo* variants, p.(Glu251Gln) in Individual 1, p.(Met255Arg) in unrelated Individuals 2 and 3, and p.(Leu262Pro) in Individual 4 are absent from gnomAD or other public genomic databases. They are located in the HLH domain of the protein, crucial for protein dimerization and highly intolerant to missense variations (Figure 1b), which indicate the importance of this region for normal protein function. All affected residues are fully evolutionarily conserved from human to zebrafish (Figure 1c). We also describe seven individuals (Individuals 5-11) with the same homozygous frameshift variant, p.(Gly74Alafs*18) in the glycine-rich region of the protein. This variant has not been reported in the homozygous state in gnomAD and, in conjunction with the strikingly similar clinical phenotype of individuals carrying this biallelic variant, strongly suggest pathogenicity.

Brain imaging scans revealed either partial (2/11) or complete (8/11) absence of the corpus callosum. In one individual, the corpus callosum was normal, but the level of penetrance for callosal agenesis across the cohort is remarkable. All ACC scans in the cohort are associated with colpocephaly, a brain anatomical feature characterized by the abnormal enlargement of the occipital horns of the lateral ventricles, and diminished white matter volume. The anterior commissure is not clearly visible in all scans available for review, but it appears hypoplastic in at least three individuals. In the scans that were available for internal review, the hippocampal commissure was absent.

8/9 individuals available for postnatal examination exhibited significant global neurodevelopmental delay and ID. Tone abnormalities are present in all individuals. Repetitive and compulsive behaviors, especially in the hands, were observed in 6/9 individuals. Oculomotor and/or vision concerns were observed in 7/9 individuals, including strabismus (5/9), disconjugate eye movements, retinal detachment, leukocoria, cortical visual impairment, and optic atrophy.

5/9 were reported to have either gastroesophageal reflux, gastrostomy tube dependence, and/or dysphagia. 4/9 unrelated individuals were reported to have a spectrum of seizure semiology or abnormal EEG.

Among the individuals with heterozygous variants, there is general limitation in language ability, typically restricted to syllables, words, or simple word combinations, and they are capable of either walking or crawling. 3/4 individuals either have a diagnosis of autism spectrum disorder or exhibit autistic traits. Additionally, 3/4 individuals reported hypotonia and the other individual reported appendicular hypertonia. Individual 4 is an outlier in terms of disease severity. Further studies are needed to explain the milder phenotype observed with this variant and one hypothesis could be mosaicism. Among individuals with the recurrent homozygous variant who were examined postnatally, all five are non-ambulatory and 4/5 are non-verbal with the other individual restricted to vocalizations. 4/5 individuals exhibit central hypotonia with appendicular spasticity and 3/5 exhibit dyskinesia and/or dystonia. Microcephaly is observed in all five individuals (<2 SD). 3/5 individuals reported dysphagia, 2/5 were dependent on gastronomy tubes for feeding, and 2/5 have scoliosis. These observations suggest that the complete loss of protein function due to the biallelic frameshift variants results in a more severe phenotype.

There have been previous reports detailing the role of BHLHE22 in central nervous system development outside of humans. In 2012, Ross et al described a major function of BHLHE22 in its ability to form a repressor complex by binding to sequence specific DNA elements and recruiting PRDM8, a transcription factor that inhibits DNA methylation, which secondarily impacts transcription. One of the important targets of the repressor complex is Cadherin-11 (CDH11), a cell-cell adhesion protein, that regulates the assembly of neural circuitry. Overall, the target genes are involved in neural development such as axonal guidance in the dorsal telencephalic neurons and the control of inhibitory synaptic interneurons in the dorsal horn. Notably, lack of bhlhe22 in the dorsal telencephalon of mice resulted in almost complete loss of the three main interhemispheric commissures in all animals tested (20/20): the corpus callosum, the hippocampal commissure, and the anterior commissure. These major axon tracts were similarly mistargeted in *prdm8 -/-* mice. Therefore, in mice lacking bhlhe22 or prdm8, neurons of the dorsal telencephalon survive, but their axons fail to reach their targets.

The brain structure findings of the *bhlhe22* knockout mice model overlap with the brain imaging results of our human cohort, providing strong evidence for the candidacy of *BHLHE22* as a candidate gene crucial for corpus callosum formation in the early fetal stages of development.

The majority of the *bhlhe22* and *prdm8*-null mice displayed elevated scratching behavior and resulted in development of skin lesions (Ross et al, 2012). We may consider the relationship to the stereotypies observed in our human cohort, with repetitive hand movements observed in 6/9 individuals. In 2008, Joshi et al showed that corticospinal motor neurons terminate prematurely along the pyramidal tract in the ventral hindbrain of *bhlh22e*-null mice, failing to reach to the spinal cord. Bhlhe22’s expression can be identified in specific layers of the developing cortical plate and subventricular zone of the brain (Brunelli et al, 2003), and the Bhlhe22 protein assists in the formation of the progenitor domains for spinal cord and brain development in chicks and mice (Skaggs et al, 2011). The gene, expressed in a high caudomedial to low rostrolateral gradient, was determined to help regulate the post-mitotic organization of area identities in cerebral development through downregulation of target genes. The somatosensory and caudal motor cortex of *bhlhe22* knockout mice revealed disorganization of the vibrissal barrels, as well as failure of corticospinal tract formation, underscoring the importance of *bhlhe22* in cortical development. The corticospinal tract, or pyramidal tract, is the main neuronal pathway that controls voluntary motor activity for the somatic motor system from the neck to distal extremities (Joshi et al, 2008). This could potentially explain the presence of pyramidal tract signs such as involuntary movements and appendicular spasticity, observed in many individuals within the cohort. A small fraction of the *bhlhe22*-null and *prdm8-null* mice aberrantly walked on their forepaws and appeared to be secondary to abnormal contraction of the hindpaws (Ross et al, 2012).

Previous studies also reported BHLHE22 involved in the downstream regulation and selected development of amacrine and bipolar neuronal subtypes during retinogenesis (Feng et al, 2006). In our cohort, oculomotor and/or vision issues are present in 7/9 individuals that were assessed. The issues include retinal detachment, bilateral leukocoria, cortical visual impairment, optic atrophy, disconjugate eye movement, esotropia, hypermetropia, myopia, and astigmatism.

In this paper, we describe for the first time, the gene *BHLHE22,* as crucial for the normal development of the human corpus callosum and other essential midline commissures. The majority of the *BHLHE22* cohort (10/11) present with ACC, partial in two individuals and complete in eight individuals. No other significant anatomical brain anomalies were observed in individuals with ACC. While generally individuals with isolated ACC have a good prognosis when compared to hypoplastic corpus callosum or ACC with additional brain malformations (Romaniello et al, 2017; Argilli and Sherr, personal communication), all individuals with ACC here present with severe clinical features, including inability to walk or talk, impaired cognition, spasticity, and/or seizures. Therefore, there should be consideration of other mechanisms affected by the disruption of normal BHLHE22 function contributing to their severe neurological phenotype.

As of today, there are hundreds of genes and syndromes linked to CCD. However, the cause of isolated ACC is solved with a genetic diagnosis least frequently amongst all CCD cases (30-40% vs 40-55%), even after trio exome sequencing is performed. Our continuous efforts to expand the understanding of the genetic causes of this disorder and to correlate phenotypes with genotypes reveal how a positive genetic diagnosis can offer valuable context regarding the clinical outcome and symptom management for families and clinicians. This is exemplified by this discovery of a previously unknown neurodevelopmental and callosal agenesis syndrome caused by recurrent biallelic and monoallelic *BHLHE22* variants.

As previously reported, BHLHE22 forms a repressor complex with PRDM8 to influence the downregulation of target genes linked to correct neural circuit formation, such as *CDH11*.

There have been previous reports of *PRDM8* and *CDH11* variants in humans with developmental delay/ID, seizures, and movements disorders. There is a provisional relationship between homozygous variants in *PRDM8* and progressive myoclonic epilepsy, ataxia, dysarthria, developmental delay, and other neurodegenerative disorders (Turnbull et al, 2012, Davarzani et al, 2022). Biallelic and heterozygous pathogenic variants in *CDH11* cause a syndrome characterized by facial dysmorphisms and ID (Harms et al, 2018, Li et al, 2021).

To our knowledge, this report demonstrates the first evidence linking a gene from the Class II bHLH superfamily of transcription factors, BHLHE22, with corpus callosum dysgenesis, psychomotor delay, and ID while genes encoding Class I bHLH proteins, such as *TCF12* and *TCF4*, have reported such association (Paumard-Hernández et al, 2015 Davis et al, 2020, Goodspeed et al, 2018, Chen et al, 2021). With the genetic and clinical phenotypes of the affected individuals described here, along with a previously reported recessive mouse model (Ross et al, 2012), we propose that pathogenic *BHLHE22* variants, both with autosomal dominant (AD) and autosomal recessive (AR) inheritance pattern, are associated with ACC, severe global developmental delay/ID, autistic features, tone and movement abnormalities, and oculomotor/vision deficits. Individual 4 does not show severe psychomotor delay or callosal anomalies, but showed young adult onset of lower limb spasticity, suggesting that the clinical phenotype might be broader, and also overlap with a hereditary spastic paraplegia phenotypes.

With additional cases in the future, it may be possible to differentiate between two distinct syndromes—one with AD inheritance and another with AR inheritance. Based on the present cohort, *BHLHE22* demonstrates a strong correlation to corpus callosum dysgenesis with severe neurodevelopmental delay and tone as well as movement abnormalities and its addition to the gene testing panel of CCD and spasticity should be considered. Future functional studies will help further define the cellular mechanism correlating the clinical symptoms with pathogenic *BHLHE22* variants.

## Data Availability

All data produced in the present work are contained in the manuscript.

## Acknowledgements

We thank the families who participated in this study. This work was supported by NIH grant R01NS058721. Sequencing and analysis of Individual 1 was provided by the Broad Center for Mendelian Genomics, funded by the National Human Genome Research Institute grants UM1HG008900, U01HG0011755 and R01HG009141. The research was also funded by the Wellcome Trust (WT093205MA, WT104033AIA), the Medical Research Council (MR/S01165X/1, MR/S005021/1, G0601943), The National Institute for Health Research University College London Hospitals Biomedical Research Centre, Rosetrees Trust, Ataxia UK, Multiple System Atrophy Trust, Brain Research United Kingdom, Sparks Great Ormond Street Hospital Charity, Muscular Dystrophy United Kingdom (MDUK), Muscular Dystrophy Association (MDA USA) and the King Baudouin Foundation. SE and HH were supported by an MRC strategic award to establish an International Centre for Genomic Medicine in Neuromuscular Diseases (ICGNMD) MR/S005021/1.

