## Supplementary figures and images for "Basic helix-loop-helix transcription factor *BHLHE22* monoallelic and biallelic variants cause a neurodevelopmental disorder with agenesis of the corpus callosum, intellectual disability, tone and movement abnormalities"

### Supplemental Figures 1 & 2

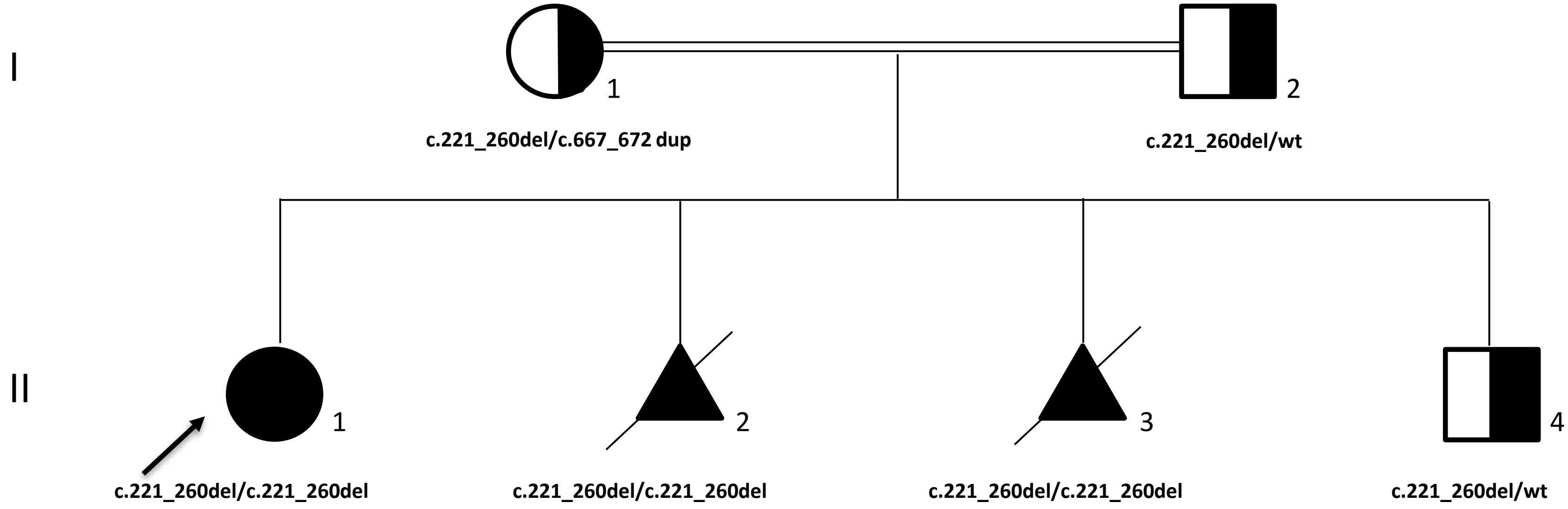

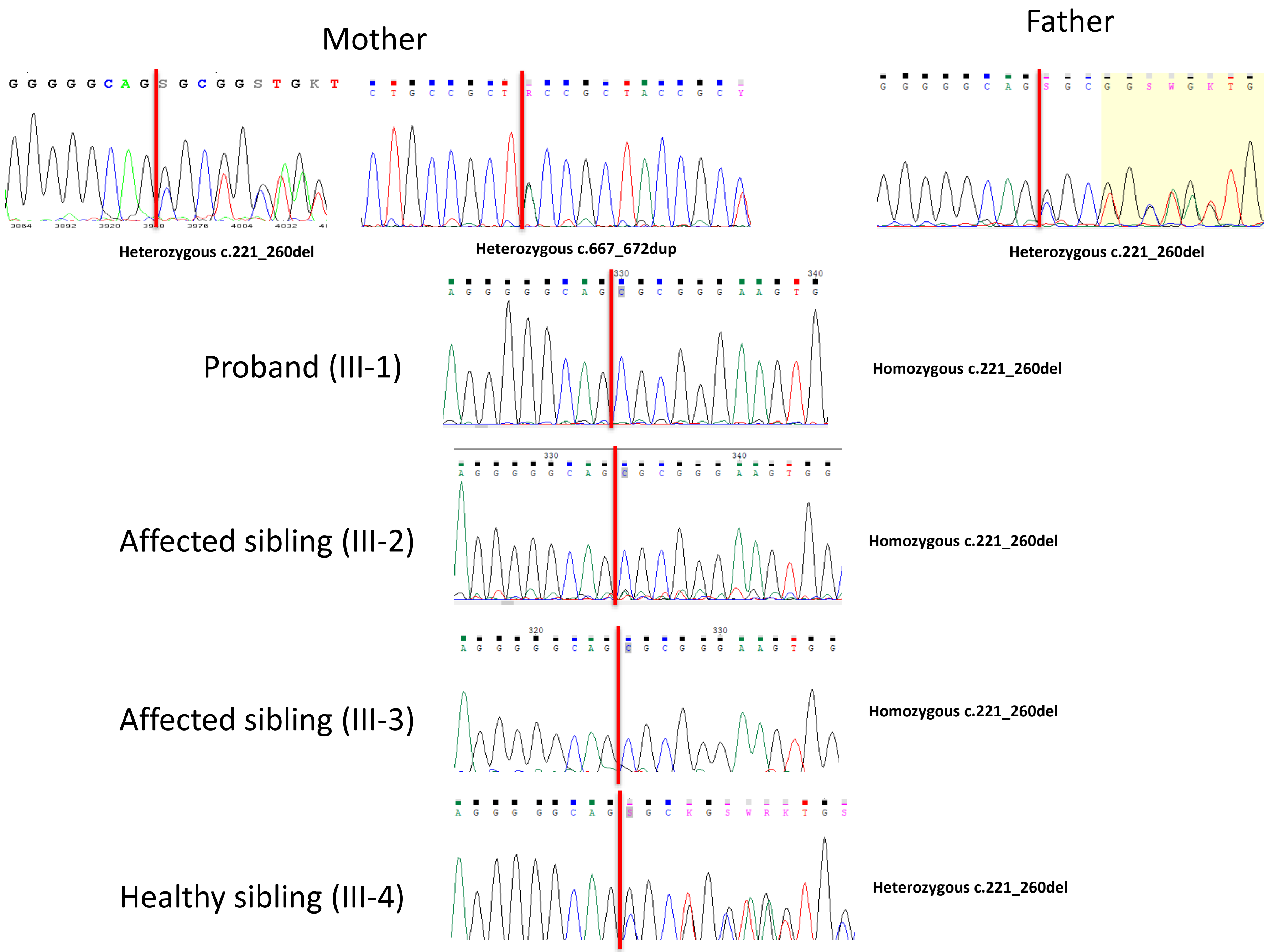
